# Antibody profiles across H5N1 and previously circulating viruses are highly dynamic and age- and imprint- independent

**DOI:** 10.64898/2026.08.26.26361396

**Authors:** Martin Beukema, Eva Vermeulen, Jacqueline de Vries-Idema, Anke Huckriede, Manas Joshi

## Abstract

The increasing incidence of H5N1 influenza virus transmission from animal species to humans has heightened concerns about an imminent H5N1 pandemic. Prior studies using recombinant hemagglutinin and neuraminidase proteins have reported age-dependent cross-reactivity to H5N1, attributed to immune imprinting from an individual’s first influenza virus exposure. However, whether this pattern holds when using whole inactivated virus (WIV), capturing antibodies against diverse viral proteins, and is stable over time remains unknown. We therefore aimed to determine whether H5N1 cross-reactivity of pre-existing antibodies to whole virus follows an age-dependent or imprinting-specific pattern, and whether this pattern is stable over a five-year period.

To this end, we measured serum antibody levels in adolescents, adults and seniors by ELISA using whole inactivated H5N1 virus as antigen rather than purified proteins. Detectable, albeit generally low, levels of H5N1-reactive antibodies were present in most individuals, irrespective of age. Comparison of antibody levels against H5N1 with those to five historical influenza virus strains revealed a consistent positive correlation between H5N1-reactive antibodies and responses to the H1N1pdm09 strain A/California/7/2009 (CA), across all age groups.

Using unbiased clustering of antibody titers against H5N1, CA, and the H3N2 strain A/Perth/16/2009 (PE), we identified seven distinct age-transcending antibody profiles. These profiles covered individuals with varying titers to all three included viruses but also identified individuals with high anti-CA levels, yet low anti-H5N1 levels and vice versa. Moreover, despite stable antibody levels over a five-year interval in the study population, individual antibody levels and profiles fluctuated considerably over this period.

Taken together, our results confirm the presence of H5N1-reactive antibodies in human sera and their association with previously circulating strains. However, they also caution against inferring antibody levels against a new strain based solely on responses to antigenically related strains and highlight the limitations of extrapolating immune status from single timepoint measurements.

## Introduction

Highly pathogenic avian influenza A(H5N1) viruses remain a persistent pandemic threat due to their continued circulation in avian reservoirs, regular bridging of the species barrier to mammalian hosts and livestock, and zoonotic transmission to humans, associated with high mortality rates (1–3). Although sustained human-to-human transmission has not yet been established, the immunological distance between H5N1 and currently circulating seasonal influenza A viruses raises concerns about the level and breadth of pre-existing population immunity (4). Importantly, such cross-reactive immunity is unlikely to be uniformly distributed across individuals, underscoring the need to identify and stratify population subgroups based on their cross-reactive immune profiles.

Several studies examining humoral immunity to H5N1 in individuals without known exposure to avian influenza have consistently reported the absence or very low prevalence of hemagglutination-inhibiting or neutralizing antibodies against H5N1 (5–7). However, these studies demonstrated the presence of cross-reactive antibodies binding to H5N1 antigens, in particular hemagglutinin (HA) and neuraminidase (NA), that might contribute to protection from severe disease by Fc-mediated effector mechanisms (5). Notably, these cross-reactive responses appeared to be strongly age-dependent, with older adults, on average, exhibiting higher levels than younger individuals (8). This age-associated pattern has been attributed to immune imprinting, where the first influenza virus encountered early in life leaves a lasting imprint on antibody repertoires and cross-reactive potential later in life (5,6,8).

Evidence for immune imprinting in the context of H5N1 has, however, largely relied on assays measuring HA- and NA-specific antibodies using recombinant proteins (5,6,8). Interestingly, an earlier study investigating antibodies binding to whole inactivated virus (WIV) preparations from five historical H1N1 and H3N2 strains did not observe age-dependent imprinting patterns (9). Instead, individuals with low and high levels of WIV-binding antibodies were distributed across all age groups. Overall, the highest antibody titers were observed against strains that had most recently circulated, even among older individuals who had first encountered influenza virus in the 1940s. Unlike HA- or NA-focused assays, WIV enables assessment of antibodies targeting the full repertoire of influenza virus proteins, including conserved internal antigens such as nucleoprotein (NP) and matrix 1 (M1) proteins (4). As such, whole-virus approaches may more accurately reveal cumulative lifetime influenza virus exposure and the resulting complexity of antibody repertoires as elicited through repeated influenza virus infections.

The age-stratified prevalence and levels of H5N1 WIV-reactive antibodies have not been investigated so far. Moreover, it is unknown whether individual and population antibody levels are stable over time. Addressing these questions is essential for refining our understanding of pre-existing humoral immunity to avian influenza and for identifying individuals at increased risk for severe disease due to low levels of cross-reactive immunity. Therefore, we quantified H5N1-cross-reactive antibody responses using H5N1 WIV and compared them with antibody levels against five influenza A strains circulating between 1940 and 2009. To this end, we used sera and antibody data from adolescents, adults, and older adults (called ‘seniors’ in the following), obtained from LifeLines, a longitudinal age-stratified cohort study running in Groningen, The Netherlands. By stratifying antibody responses by age and correlating H5N1 reactivity with strain-specific antibody profiles, we aimed to determine whether H5N1 cross-reactivity at the whole-virus level follows an age-dependent or imprinting-specific pattern. Exploitation of sera taken at two timepoints 5 years apart enabled us to investigate the persistence of the observed profiles.

Our results demonstrate the presence of detectable but low levels of H5N1 WIV-reactive antibodies in most of the investigated sera, with only minor age-related differences. In all age groups, H5N1 antibody levels showed a strong and significant correlation with the antibody levels to the recently circulating H1N1pdm09 influenza virus strain. Nevertheless, the presence of H1N1pdm09-reactive antibodies was neither a prerequisite nor a guarantee for the presence of H5N1 WIV-reactive antibodies. Overall, we found age-transcending, yet dynamic, antibody profiles across the panel of influenza virus strains, likely originating from complex individual-specific infection and vaccination histories. Taken together, our findings add vital knowledge by refining our understanding of pre-existing humoral immunity to avian influenza.

## Material & Methods

### Study setup

The study design was similar to that of our previous studies (4,9). In brief, human serum samples were collected from participants enrolled in the LifeLines Biobank, a large multigenerational, population-based cohort study consisting of over 167,000 participants from the northern Netherlands. Participants were stratified into three age categories: adolescents (17-18 years old, 15 males and 45 females), adults (37-41 years old, 30 males and 30 females), and elderly (62-67 years old, 30 males and 30 females). All age categories had equal sample size, n=60. All recruited participants did not self-report any pre-existing medical conditions, such as cancer, diabetes, or asthma. Blood samples were collected from 180 individuals at two assessment timepoints: Assessment point 1 (A1) and Assessment point 2 (A2). For adults and elderly, A1 and A2 were in 2009 and 2014, respectively; for adolescents, A1 and A2 were in 2011-12 and 2015-17, respectively [see Figure 1A].

**Figure 1.**
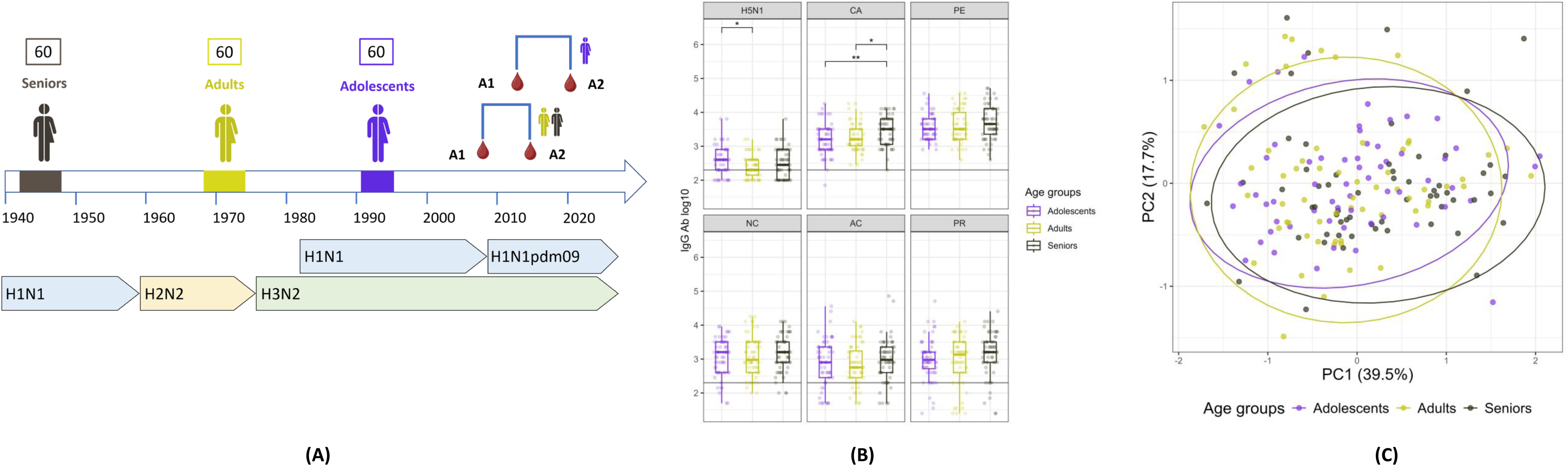
Influenza virus antibody profiles among age groups (A) Schematic representation of the study setup: Depicted are the birth periods of the senior, adult and adolescent participants, the assessment timepoints A1 and A2 for the different age groups, and the influenza A virus subtypes circulating over time. (B) Distribution of the antibody titers against the six studied virus strains across age groups for A1 samples. Antibody titers were normalized using a logarithmic scale. Pairwise titer distributions were compared using Tukey’s HSD test, significance values are indicated as: p < 0.05 (*), p < 0.01 (**), and p < 0.001 (***). Detection limits of antibody titers were set at 200. PR = A/Puerto Rico/8/1934 (H1N1), PE = A/Perth/16/2009 (H3N2), NC = A/New Caledonia/20/1999 (H1N1), AC = A/Aichi/1/1968 (H3N2), and CA = A/California/07/2009 (H1N1). (C) Principal component analysis (PCA) of the overall antibody titer makeup against the six studied strains for A1.

### Preparation of Whole Inactivated Virus of the H5N1 influenza virus

The H5N1 influenza virus strain NIBRG-23 (National Institute of Biological Standards and Controls, Potters Bar, United Kingdom) was generated by reverse genetics and contains gene segments from A/turkey/Turkey/1/2005 (H5N1) and A/Puerto Rico/8/34 (H1N1). Virus stocks were propagated in Madin–Darby canine kidney (MDCK) cells. Whole inactivated virus (WIV) was produced by incubating harvested and purified virus with 0.1% (v/v) β-propiolactone (Acros Organics, Geel, Belgium) overnight at 4 °C under continuous rotation. The inactivated material was dialyzed overnight at 4 °C against HEPES-buffered saline (Thermo Fisher Scientific, Bleiswijk, The Netherlands) to remove residual β-propiolactone. Complete inactivation was verified by inoculating MDCK cells and assessing for viral replication using a hemagglutination assay (10).

### Influenza-specific IgG enzyme-linked immunosorbent assay (ELISA)

Influenza-specific IgG responses were quantified using enzyme-linked immunosorbent assays (ELISAs) targeting whole inactivated virus (WIV), following established protocols (4,11). ELISA plates were coated with WIV at 0.3 µg per well. Serum samples were pre-diluted 1:200 and applied in two-fold serial dilutions. Bound IgG was detected using horseradish peroxidase-conjugated goat anti-human IgG (SouthernBiotech, cat. 2040-05). Color development was performed with O-phenylenediamine dihydrochloride (Sigma-Aldrich, USA), and absorbance was measured at 492 nm. Data of influenza-specific IgG to WIV derived from A/Puerto Rico/8/1934 (H1N1; PR), A/Perth/16/2009 (H3N2; PE), A/New Caledonia/20/1999 (H1N1; NC), A/Aichi/1/1968 (H3N2; AC), and A/California/07/2009 (H1N1; CA) were obtained from a previous study by Sicca et al. (9).

### Statistical analyses and visualization

All analyses and visualizations were performed using the R programming language (12) (version 4.6.1). Antibody titers against each strain were log-transformed prior to analysis. Pairwise comparisons between groups were performed using Tukey’s honestly significant difference (HSD) test, with statistical significance defined as p < 0.05 (*), p < 0.01 (**), and p < 0.001 (***). The antibody titer detection limit for all strains was set at 200. Principal component analysis (PCA) was performed using the *prcomp*function from the *stats* package (version 4.6.1). Correlation analyses were performed using the *cor.test* function from the *stats* package (version 4.6.1) with the Pearson correlation method. Associations between H5N1 antibody titers and antibody titers against previously circulating strains were assessed using linear regression models, with H5N1 titers as the dependent variable and titers against other strains as independent variables. Linear models were fitted using the *lm* function from the *stats* package (version 4.6.1). Visualizations were performed using the *ggplot2* package (version 4.0.3). The analyses and visualization scripts are stored on publicly available GitHub repository - github.com/Manaswwm/H5N1_cross_reactivity.

### Clustering

To group individuals based exclusively on their antibody titer profiles across strains, an unbiased clustering approach was applied. For each strain, antibody titer quartiles were calculated, and individuals were assigned to one of four categories: Q1 (titer below the first quartile), Q2 (titer between the first quartile and median), Q3 (titer between the median and third quartile), and Q4 (titer above the third quartile). Individuals were assigned a strain-specific score corresponding to their quartile category, resulting in a composite antibody profile used for clustering. Based on these quartile-based antibody profiles, individuals were clustered using an unsupervised approach. Pairwise distances were calculated using the *dist* function from the *stats* package (version 4.6.1) with the Manhattan distance metric. The optimal number of clusters was determined using the *fviz_nbclust* function with method *silhouette* from the *factoextra* package (version 2.2.0).

## Results

### Similar antibody titers against WIV of H5N1 and other influenza virus strains across all age groups

To identify cross-reactive antibody signatures associated with H5N1, we focused on a small subset of sera and serological data of adolescents, adults, and seniors (n=60 per subgroup) from the large longitudinal population-based cohort study LifeLines (see M&M for further details). The first set of sera (assessment 1 (A1)) had been collected in 2009 (adults, seniors) and in 2011/12 (adolescents), respectively, the second set (assessment 2 (A2)) in 2014 and 2015/16 [see Figure 1A].

Using the A1 samples, we first quantified H5N1-binding antibodies by enzyme-linked immunosorbent assay (ELISA), employing plates coated with H5N1 WIV. The majority of the included sera harbored measurable amounts of H5N1-binding antibodies, with the prevalence being highest in adolescents (83.7%), followed closely by adults (78.3%) and seniors (73.3%). Adolescents also displayed the highest H5N1 antibody levels, although the differences from the levels in other age groups were small and significant only for the adult group [Figure 1B].

In the same sera used for the H5N1 ELISAs, we had earlier determined antibody titers against WIV preparations of five previously circulating influenza virus strains: A/Puerto Rico/8/34 H1N1 (PR), A/Aichi/1/68 H3N2 (AC), A/New Caledonia/20/99 H1N1 (NC), A/Perth/16/2009 H3N2 (PE), and A/California/7/2009 H1N1pdm09 (CA) (Sicca et al 2022). Across all age groups, the H5N1-reactive antibody levels were significantly lower than those to either of the previously circulating strains [Supplementary Figure 1]. Yet, despite strain-dependent differences in the total antibody levels, we noted that the overall age group-specific antibody profiles across all six strains were largely similar [Figure 1B], as also demonstrated by the absence of distinct age-related clusters in a principal component analysis (PCA) involving the antibody levels to all strains [Figure 1C]. This result reflects considerable variation within each age group rather than between age groups.

### H5N1 antibody titers are most strongly associated with CA antibody titers

Next, to identify age group-specific signatures reflective of cross-reactivity to H5N1, we performed titer-based correlation analyses [Figure 2A]. We note that H5N1-reactive antibody levels in all age groups exhibited significant positive correlation with the antibody levels to the recently circulating H1N1pdm09 strain CA. Interestingly, H5N1-reactive antibody titers of adolescents exhibited a multi-strain cross-correlation pattern (significant positive correlation with antibody titers to 4/5 strains), in contrast to antibody titers in adults and seniors (significant positive correlation to antibody titers to 1/5 strains).

**Figure 2.**
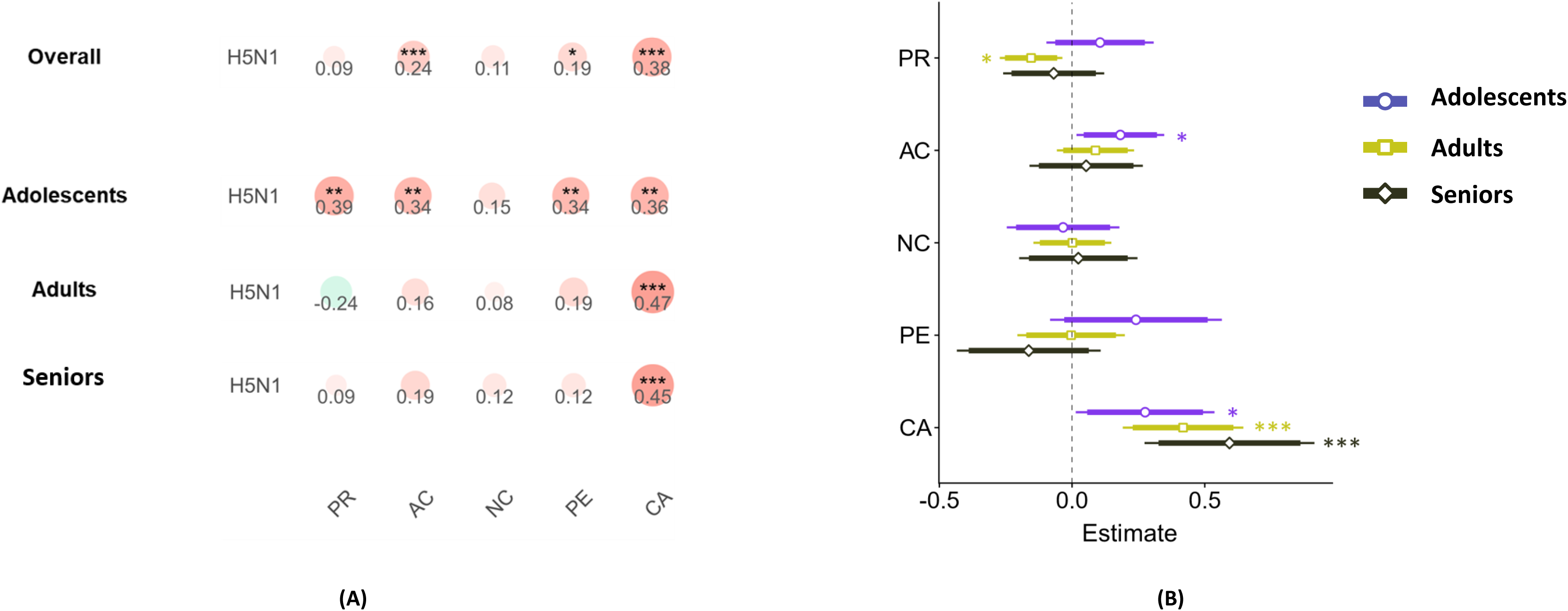
Associations between antibody titers to previously circulating influenza virus strains and the H5N1 strain. (A) Correlations of the titers against the H5N1 strains with the titers against the five previously circulating strains: overall and faceted by age categories, at assessment point A1. Correlations were performed using the Pearson method, and significance values are indicated as: p < 0.05 (*), p < 0.01 (**), and p < 0.001 (***). Numbers in circles indicate the correlation coefficients. PR = A/Puerto Rico/8/1934 (H1N1), PE = A/Perth/16/2009 (H3N2), NC = A/New Caledonia/20/1999 (H1N1), AC = A/Aichi/1/1968 (H3N2), and CA = A/California/07/2009 (H1N1pdm09). (B) Linear modelling to highlight the associations between H5N1 and the previously circulating strains based on their respective antibody titers at assessment point A1. Here, H5N1 titers are dependent variables, whereas those against the five previously circulating strains are independent variables. Estimate (x- axis) represents the putative change in H5N1 titer associated with a one-unit increase in titer against the corresponding previously circulating strain.

To further supplement this, we applied a linear modeling approach, using antibody titers against the five previously circulating strains as independent variables and titers against H5N1 as the dependent variable [Figure 2B]. In line with the correlation analyses, we note that only the titers against the CA strain showed a significant positive association with H5N1-reactive titers across all age groups. Only in adolescents, a (trend towards a) positive correlation was also found for older virus strains. Taken together, these findings suggest that the immune memory from the recent CA strain may play a key role in conferring potential cross-reactive functionality against the H5N1 strain.

### Age-transcending antibody profiles across different influenza virus strains

To get further insight into age-related antibody profiles, we shortlisted the two recently circulating strains (CA and PE) besides H5N1. Analyzing the antibody profiles for the three strains by PCA, we noted that adolescents form a tighter cluster, indicating reduced within-group variation when compared to the more dispersed older groups [Figure 3A]. This suggests a higher degree of antibody profile similarity for these strains, possibly due to a smaller antibody repertoire, among adolescents than among the other age groups. However, similar to the PCA analysis mentioned above [Figure 1B], we were unable to detect distinct, non-overlapping clusters specific to each age group, implying a significant overlap in terms of the antibody profile amongst the age groups.

**Figure 3.**
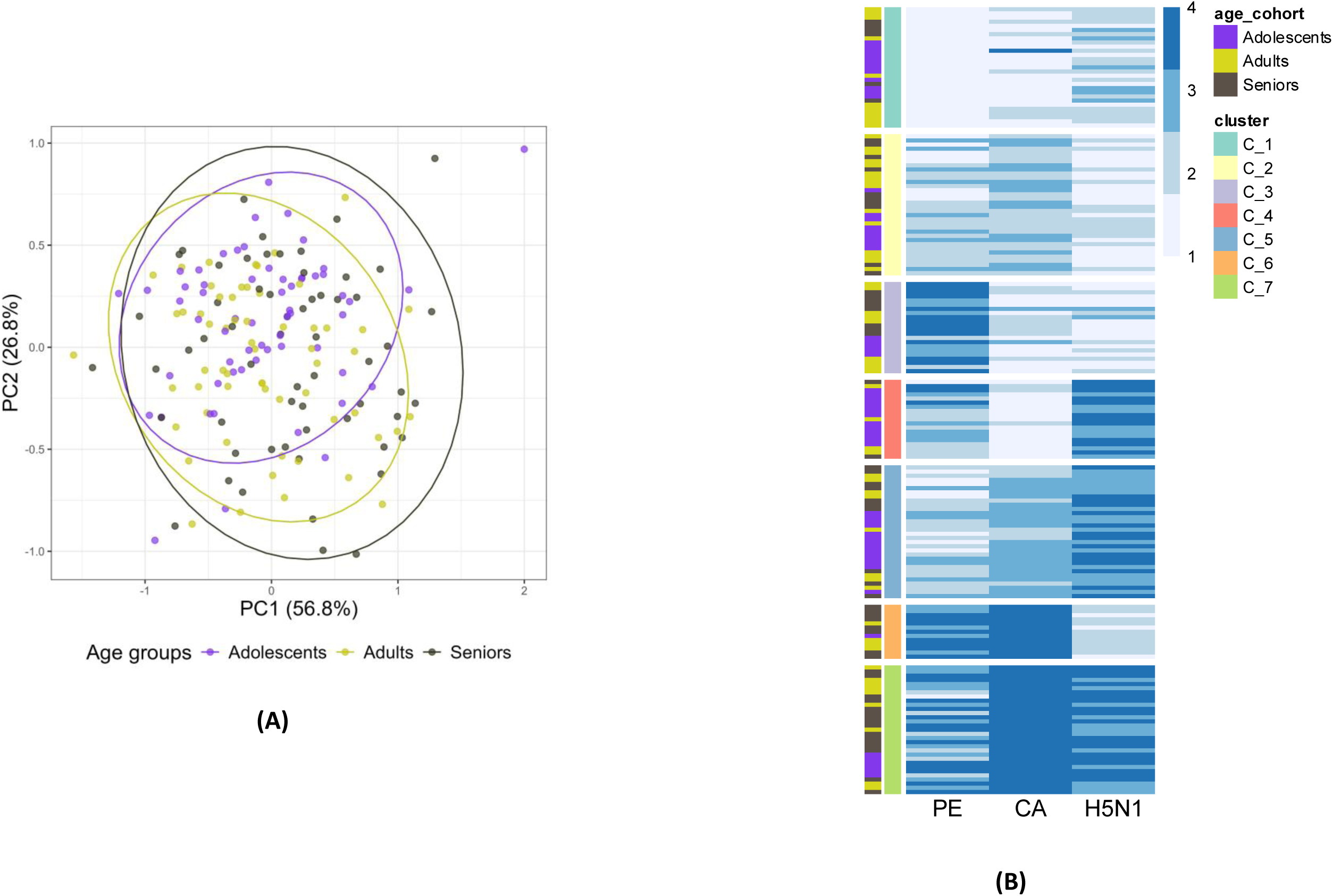
Age-transcending antibody profiles against H5N1, CA, and PE at assessment point A1. (A) PCA of the overall antibody titer makeup against the two strains that circulated around the A1 sampling moment (PE and CA). (B) Unbiased clustering of individuals at A1 based on their respective quartile-based scores against PE, CA and H5N1. Individuals are depicted as rows and the extreme left color bar is indicative of their respective age groups. Strains are depicted as columns and the individual-specific quartile scores are depicted with a four-color palette, with lighter shades indicating lower score (comparatively low titers) and darker shades indicating higher score (comparatively high titers).

Hence, we next performed unbiased clustering to identify possible age group-transcending antibody clusters [Figure 3B]. To this end, we used a quartile-based approach (see Materials and Methods) to score individuals based on their per-strain titers. In brief, for each strain, individual antibody titers were stratified into four quartiles, assigning scores from 1 (lowest, Quartile 1) to 4 (highest, Quartile 4). Using this stratification, we were able to identify seven clusters in total, including two extreme clusters: Cluster 1 (C_1) – overall low to low-medium titers against the three strains, and Cluster 7 (C_7) – overall medium to medium-high titers against the three strains. Importantly, all three age groups were represented in both the extreme clusters, albeit in different numbers [Supplementary Table 1]. These data indicate that the antibody clusters are not exclusively influenced by age but also by individual-specific factors, including personal exposure history.

In addition to the extreme clusters, we also identified clusters representing heterogeneous signatures against the three strains. Regarding H5N1-reactive antibodies, clusters C_4 and C_6 are particularly interesting. Cluster C_4 harbors individuals with moderate PE-reactive antibodies, very low CA-reactive antibodies but high levels of H5N1-reactive antibodies. On the other hand, Cluster C_6 captures individuals showing medium to high titers for PE, high titers for CA, and low to medium titers for H5N1. Given the phylogenetic proximity of CA (H1N1) and H5N1, both belonging to Group 1 influenza viruses, these clusters showcase unique and unexpected combinations of strain-specific signatures. Thus, despite the strong correlation between H5N1- and CA-reactive antibody titers in the population-based analysis, on an individual level, CA-reactive antibody responses were neither required nor predictive for H5N1-reactive responses.

### Individual antibody profiles are dynamic over time

Following the in-depth analyses of the sera taken at A1, we next focused on the A2 sera, collected 5 years later, to evaluate whether the signals detected in the A1 samples were stable over time. In the A2 sera, we found that the distribution patterns of the titers for the six strains across the three age groups were largely similar to those observed at A1 [Figure 4A]. Specifically, titer distributions showed comparable trends across age groups, with only one significant difference noted—seniors vs adults for CA (Tukey’s HSD, adjusted p-value < 0.05). The H5N1 titers remained relatively similar across age groups at A1 and A2, suggesting limited age group-level variation [Supplementary Figure 2]. In addition, similar to the A1 sera, the titers to H5N1 were lower as compared to the titers to the other five strains [Supplementary Figure 3], and we could not find age group-specific clusters [Supplementary Figure 4].

**Figure 4.**
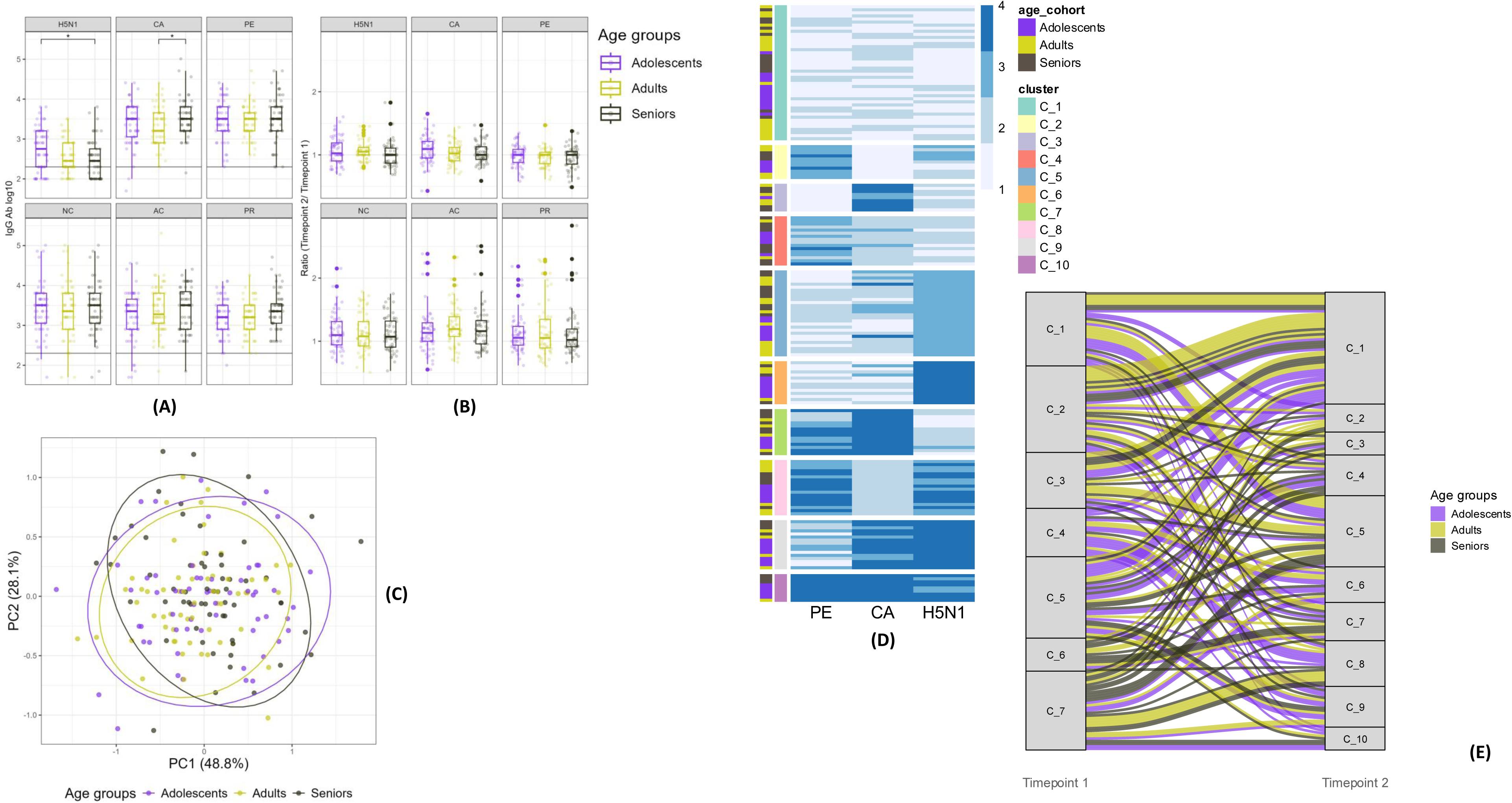
Temporal dynamics of an individual’s antibody titers and age-transcending profiles between assessment points A1 and A2. (A) Distribution of titers against the six studied strains across age groups for assessment point A2. Like for A1 samples, titers were normalized using a logarithmic scale and pairwise titer distributions were compared using Tukey’s HSD test. (B) Changes in the antibody titers between A1 and A2 depicted using a ratio metric – A2 titers/A1 titers. Here, ratio > 1: comparatively high titers at assessment point 2, ratio < 1: comparatively high titers at assessment point 1, ratio = 1: similar titers at assessment point 1 and assessment point 2. Distributions were compared using Tukey’s HSD test. (C) PCA of the overall antibody titer makeup against the recent two circulating strains (PE and CA) and H5N1 for assessment point A2. (D) Unbiased clustering of individuals at assessment point 2 based on their respective quartile-based scores against PE, CA and H5N1. Similar to Figure 3(B), individuals are depicted as rows and their respective age groups indicated on the extreme left color bar. Strains are depicted as columns and individual-specific quartile scores are depicted with four-color palette, with lighter shades indicating lower score (comparatively low titers) and darker shades indicating higher score (comparatively higher titers). (E) Alluvial plots indicating the overlaps of the clusters at assessment point 1 and assessment point 2. Individuals are indicated as rows and the ribbon colors are indicative of their respective age groups.

To illustrate the changes in titer between A2 and A1, we calculated the ratio of titers at these assessment points (A2/A1) for all individual sera. This analysis revealed substantial individual immune variability over time, as indicated by the wide distribution of ratios across all strains, with no distinct age group-related trends [Figure 4B].

Similar to A1, we next focused on the recent two strains (PE and CA) besides H5N1. Here, we could not detect distinct age-specific clusters, indicating high variability [Figure 4C]. Next, we performed the quartile-based cluster analysis as done for the A1 samples on the A2 samples and compared the individuals’ cluster allocations for the two analyses. For the A2 samples, unsupervised clustering of the quartile stratification for PE, CA, and H5N1-reactive antibody levels resulted in the identification of 10 clusters instead of the 7 clusters for the A1 quartiles [Figure 4D]. Comparing the cluster allocations of individuals between A1 and A2 revealed a substantial flow, reflecting both increases and decreases in quartile scores [Figure 4E]. These results further indicate that antibody titers, though relatively stable over time on the population level, can vary substantially on an individual level.

## Discussion

The current study aimed to determine whether H5N1 cross-reactivity of pre-existing antibodies to whole-virus follows an age-dependent or imprinting-specific pattern. Our data demonstrate that H5N1 WIV-reactive antibodies are widespread in the general population, with most adolescents, adults, and seniors showing detectable, albeit low, titers. Unlike previous reports describing a clear age-specific pattern in H5N1 cross-reactivity attributed to immune imprinting (5–7), we found only modest differences between age groups, and no distinct age-specific antibody signatures emerged from unsupervised clustering. Instead, H5N1 WIV cross-reactivity was strongly associated with antibody levels against the most recently circulating CA strain, across all age groups. Unbiased clustering further revealed that despite this strong correlation between H5N1- and CA-reactive antibody titers in the population-based analysis, CA-reactive antibody responses were neither required nor predictive for H5N1-reactive responses at the individual level. Finally, antibody titers and cluster association shifted considerably over a five-year period, indicating that these cross-reactive antibody profiles to H5N1 are dynamic rather than fixed traits.

A particularly consistent finding across all three age groups was a strong positive association between H5N1-reactive titers and antibody levels against CA, the most recently circulating H1N1pdm09 strain, as noted through correlation- and multivariate linear model-based analyses. This is compatible with the idea that recent exposure, whether through infection or vaccination, boosts a pool of broadly reactive antibodies (9). These antibodies may engage conserved epitopes shared between CA and H5N1, potentially through back-boosting of pre-existing cross-reactive B cell clones (13). Moreover, our findings also suggest that population immunity to H5N1 WIV is not so much shaped by the first virus encountered in childhood but rather by the most recent antigenic exposure, as was found earlier (9). This is consistent with an antigenic-seniority-like response (14,15), operating at the level of whole-virus binding antibody responses, as observed for neutralizing antibodies.

Building on this population-level association, the unbiased, quartile-based clustering approach of individual antibody responses revealed seven age-transcending antibody profiles, underscoring that H5N1 cross-reactivity to whole-virus is shaped by individual exposure and vaccination histories rather than age category per se. Even the two extreme clusters, C_1 (low across strains) and C_7 (high across strains), contained representatives of all three age groups, and their compositions only partially reflected age (e.g., an overrepresentation of adolescents in C_1). Age-independent response patterns have similarly been reported using unbiased, quartile-based clustering approaches for multiple vaccine responses (16) and for adaptive immune responses to SARS-CoV-2 (17). Moreover, although H5N1- and CA-reactive antibodies were strongly associated at the population level, this association was weak at the individual level within clusters C_4 and C_6. These findings indicate that, at an individual level, CA-reactive antibodies are not predictive of H5N1-reactive antibodies despite the phylogenetic relationship between the two strains. Together, this unbiased clustering approach enabled age-transcending categorization of individuals based solely on their immune response patterns, disentangling population-level correlation from individual-level variation (16)(17).

Another observation of this study is the intra-individual variability in antibody titers over the five-year interval between assessments. Although the overall population-level distributions and the absence of age-specific clustering were reproduced at the second assessment point, individual titer ratios varied widely and without age-related trends, resulting in altered quartile-based clusters. Most notably, the strong association between CA and H5N1 titers at the population level, a central finding at the first timepoint, weakened over time. Intra-individual variability in antibody responses over the five years may be caused by multiple factors, including differences in antibody waning, frequency and type of influenza virus exposure, level and type of pre-existing immunity (9,18,19). These observations caution for interpreting single cross-sectional serology snapshots as stable indicators of an individual’s, or even a population’s, cross-reactive potential against H5N1. Instead, our findings highlight the need for longitudinal sampling when using pre-existing antibody profiles to estimate pandemic risk or to guide vaccination strategies.

Several limitations should be considered when interpreting these findings. First, WIV ELISA titers reflect antibody binding rather than neutralizing or Fc-mediated effector function. Therefore, it remains unclear to what extent the H5N1-reactive antibodies identified here confer functional protection. Second, our sample size, while adequate for the exploratory clustering approach used, was modest (n=60 per age group). Furthermore, the history of influenza infection and vaccination of these individuals is undocumented. Third, the panel of previously circulating strains, spanning 1940-2009, does not capture more recent seasonal drift variants or the recent highly pathogenic H5N1 clades. A more contemporary or broader strain panel could reveal associations more relevant for the current situation. Fourth, the observed correlations between titers against H5N1 and the previously circulating strains are consistent with the presence of shared immune recognition but do not establish the underlying causal relationship. Functional studies assessing antibody cross-reactivity and neutralizing capacity can further cement the biological significance of the observed association patterns. Finally, the quartile-based clustering approach allowed for an unbiased, data-driven stratification. However, it imposes discrete boundaries on the input data, which may not fully capture the complexity of individual antibody landscapes.

Collectively, our findings confirm that cross-reactive antibodies against whole-virus H5N1 are common in the general population and are most closely tied to antibody levels against the most recently circulated seasonal influenza strain, irrespective of age. However, the existence of age-transcending, individual-specific antibody clusters, the partial dissociation between CA and H5N1 reactivity in some individuals, and the marked temporal dynamism of these responses together indicate that pre-existing humoral immunity to whole-virus H5N1 cannot be reliably predicted from age or from a single antibody measurement alone. Future work incorporating functional antibody assays, fine epitope mapping, vaccination and infection history, and repeated longitudinal sampling will be essential to translate these serological signatures into meaningful estimates of individual and population-level protection against a potential H5N1 pandemic.

## Data Availability

All data produced in the present study are available upon reasonable request to the authors

## Acknowledgements

The Lifelines Biobank initiative has been made possible by subsidy from the Dutch Ministry of Health, Welfare and Sport, the Dutch Ministry of Economic Affairs, the University Medical Center Groningen, University Groningen, and the Northern Provinces of the Netherlands. The authors wish to acknowledge the services of the Lifelines Cohort Study, the contributing research centres delivering data to Lifelines, and all the study participants. The authors would like to thank Federica Sicca for excellent support in data collection.

## Supplementary Figures

Supplementary Figure 1: Comparison of the titers against H5N1 to the previously circulating five strains at A1. Distributions were compared pairwise (H5N1 vs all) using Tukey’s HSD (p < 0.05 (*), p < 0.01 (**), and p < 0.001 (***)). Detection limit of the antibody titers was set at 200.

Supplementary Figure 2: Comparison of the H5N1 titers at the two assessment points (AP1 and AP2) for the three age categories. Distributions were compared using Tukey’s HSD (p > 0.05 = ns). Detection limit of the antibody titers was set at 200.

Supplementary Figure 3: Comparison of the titers against H5N1 to those against the five previously circulating strains at A2. Similar to the comparisons of A1 titers, distributions were compared pairwise (H5N1 versus all) using Tukey’s HSD.

Supplementary Figure 4. Principal component analysis (PCA) analysis of the overall antibody titer makeup against the six studied strains for A2

## Supplementary Tables

Supplementary Table 1: Age category-specific counts of individuals within the unbiased clusters obtained at assessment point 1 [see Figure 3(B)].

