## Supplementary Figures for "Antibody profiles across H5N1 and previously circulating viruses are highly dynamic and age- and imprint- independent"

### Slide 1
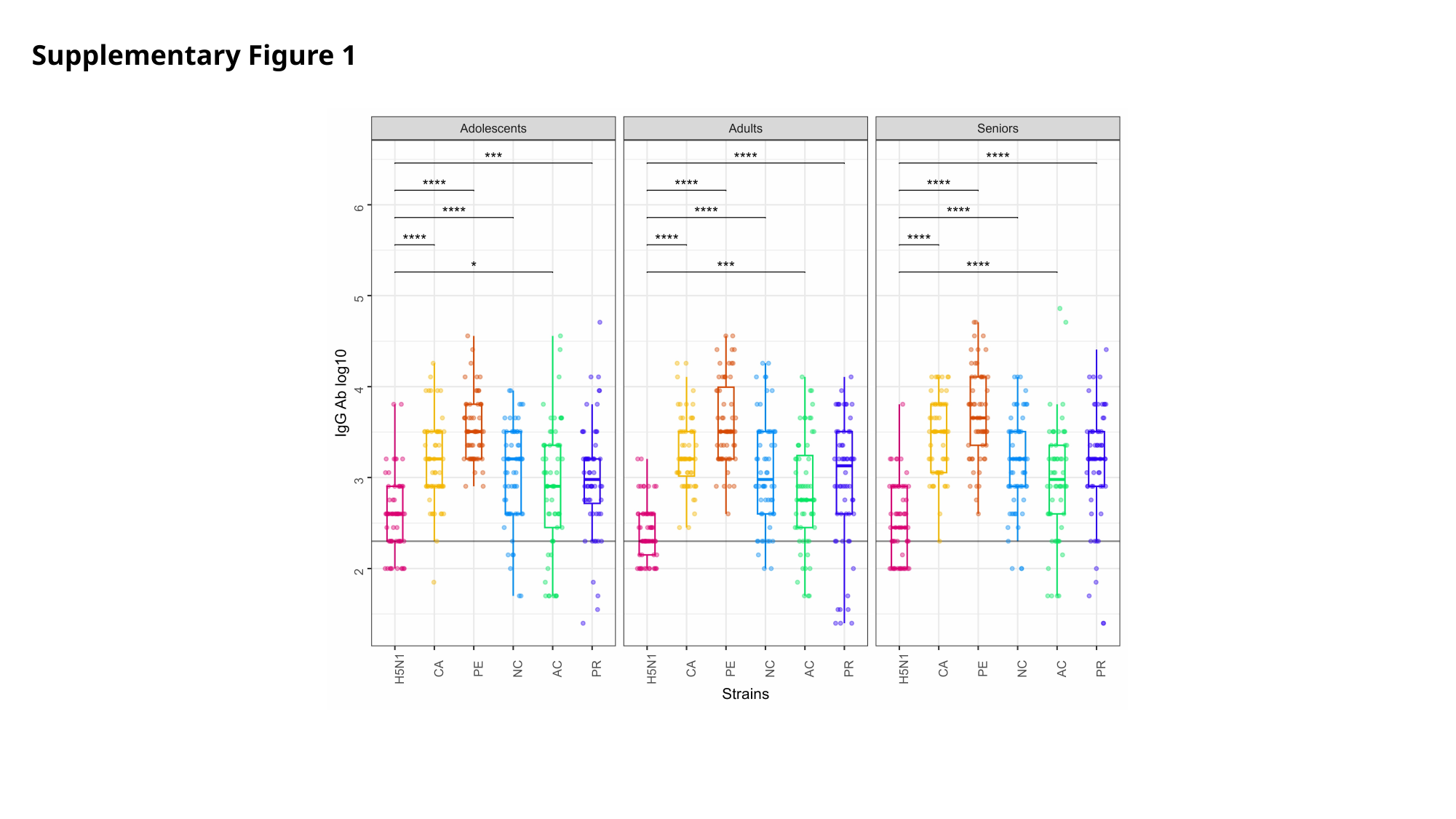

Supplementary Figure 1

### Slide 2
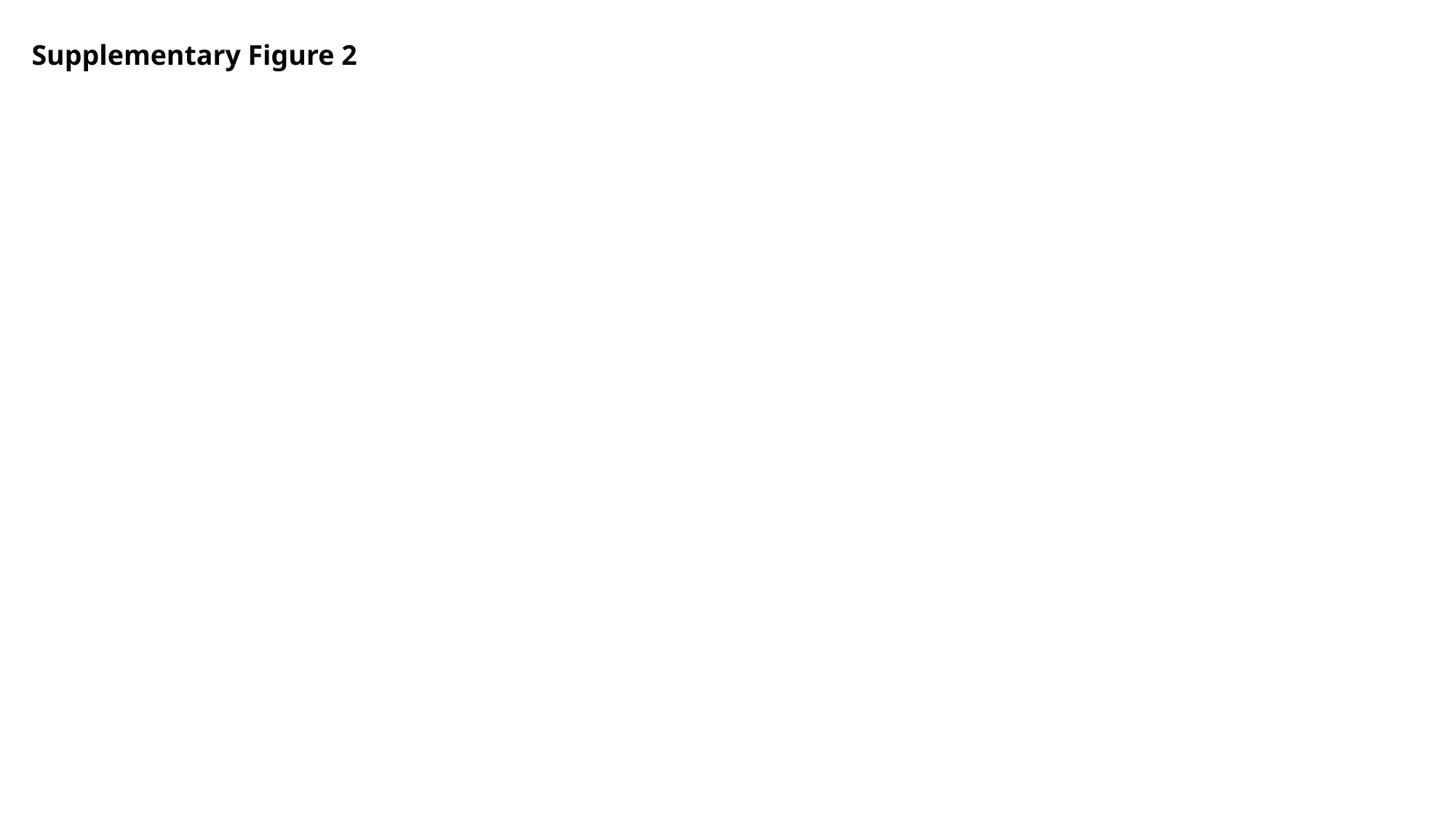

Supplementary Figure 2

### Slide 3
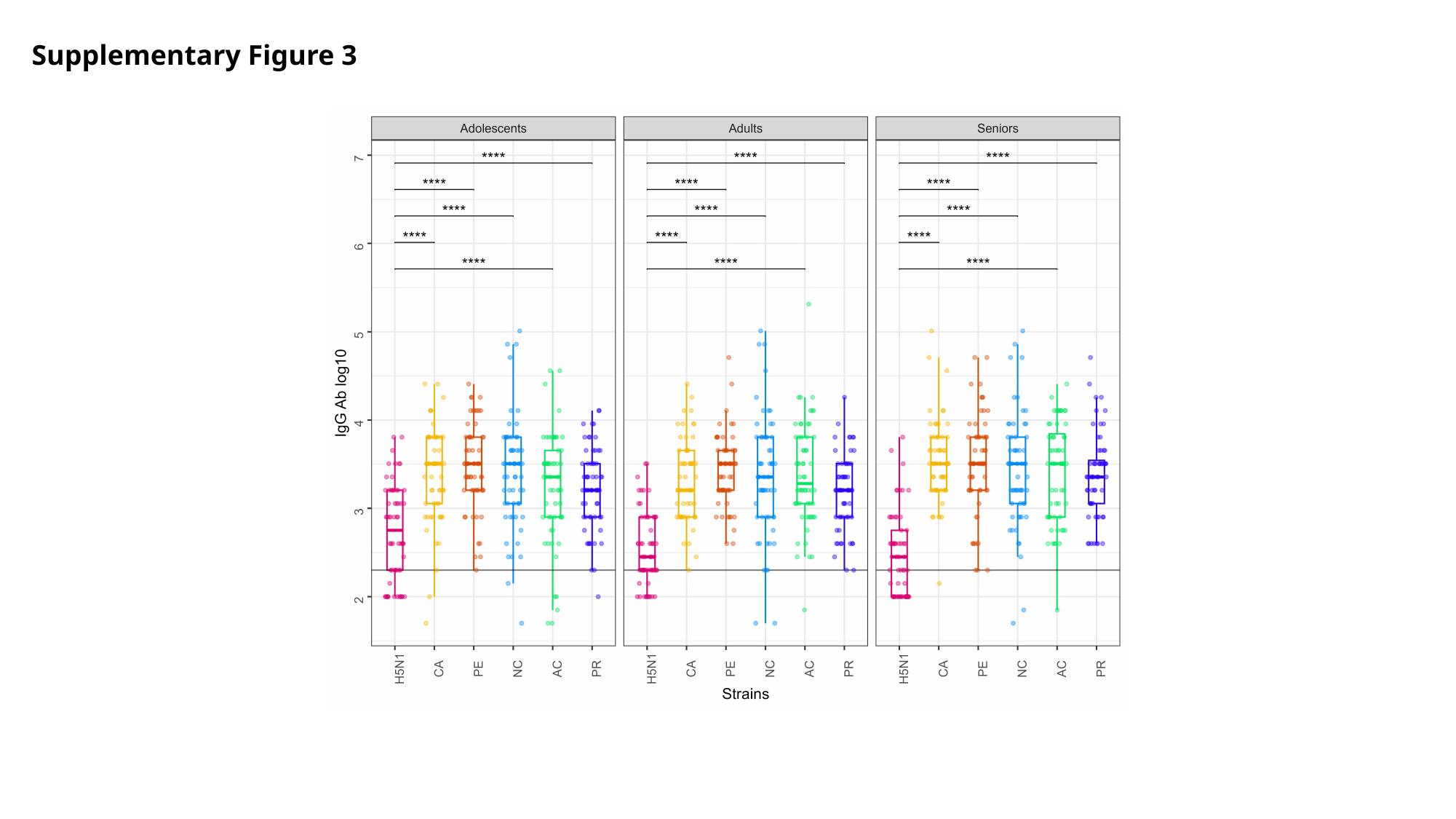

Supplementary Figure 3

### Slide 4
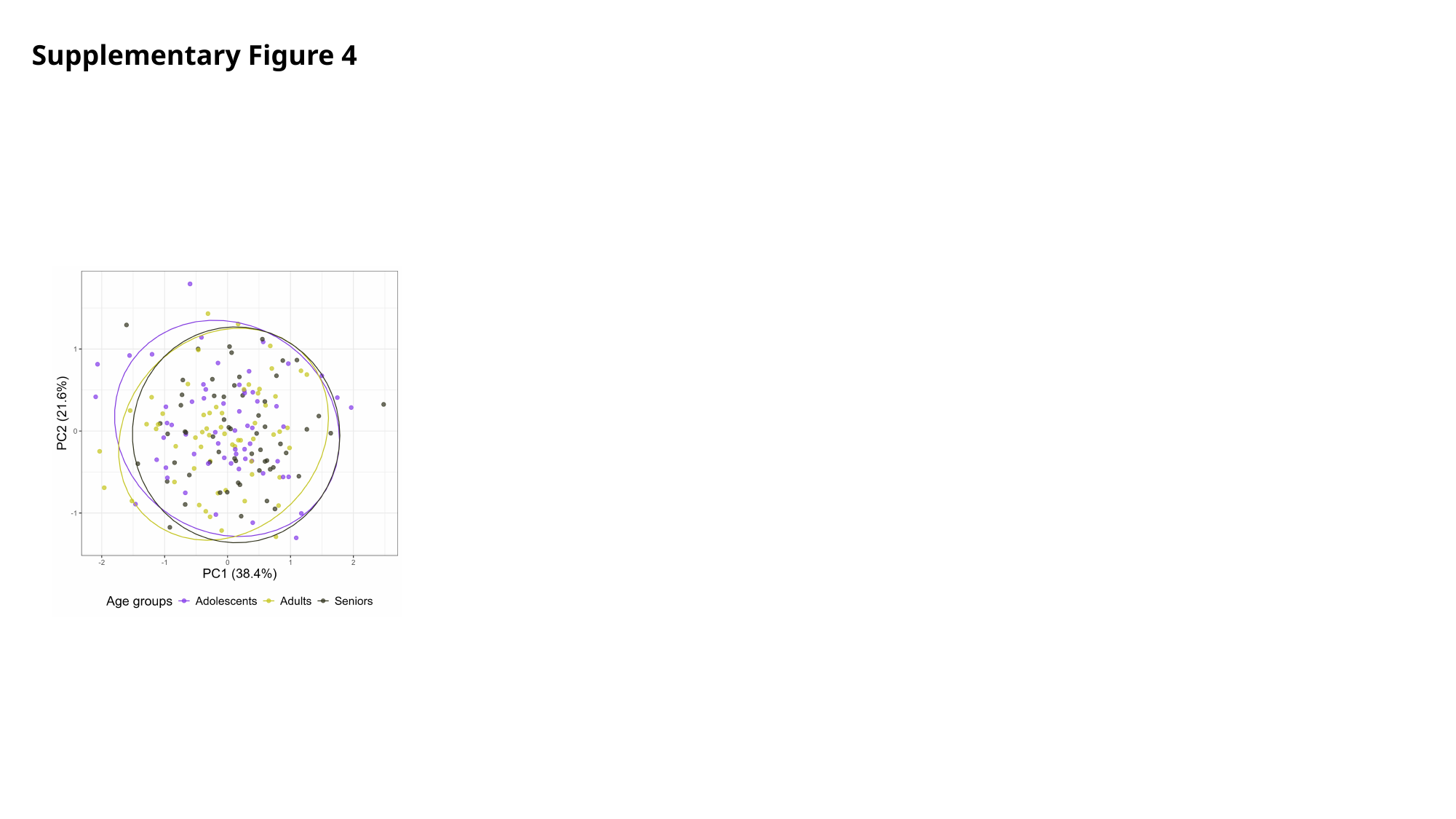

Supplementary Figure 4
